# Burden and factors associated with hospitalization among patients with chronic Diseases at a tertiary Hospital in Wolaita Zone, South Ethiopia: A Cross-sectional study

**DOI:** 10.64898/2026.10.14.26365506

**Authors:** Hailu Chare Koyra, Yalemzerf Worku Belete, Zewde Zema Kanche, Zinabu Desalegn, Mengistu Meskele Koyra

## Abstract

**Background:** Chronic diseases are the leading causes of illness, disability, and death worldwide, with a disproportionate burden in low- and middle-income countries. Hospitalization is a critical outcome in chronic disease management, reflecting disease severity, treatment failure, and healthcare utilization. However, data on hospitalization burden and its associated factors among chronic disease patients in Ethiopia, are limited. This study aimed to determine the rate of hospitalization and its associated factors among patients with chronic diseases attending a tertiary hospital in Wolaita Zone, South Ethiopia.

**Methods:** A facility-based cross-sectional study was conducted among 321 patients with chronic diseases attending the chronic follow-up clinic at Wolaita Sodo University Comprehensive Specialized Hospital from November to December 2022. Consecutive recruitment was used, with unique medical record numbers tracked to prevent double recruitment. Data were collected through structured questionnaires and medical record review. Negative binomial regression was used to identify factors associated with hospitalization rate, with incidence rate ratios (IRR) and 95% confidence intervals (CI) reported. A p-value < 0.05 was considered statistically significant.

**Results:** Of the 321 participants, 66% reported at least one hospitalization while on treatment, with a mean of 1.70 ± 1.81 hospitalizations per patient. Drug discontinuation was the most frequently documented reason for hospitalization (39.5%), followed by disease complications (22.3%) and infections (11.7%). Multimorbidity was prevalent, with the majority (59.8%) had 2–3 conditions, and a mean of 2.40 ± 1.02 chronic diseases per patient. Male sex was associated with a higher hospitalization rate, while regular physical exercise was associated with a lower number of hospitalizations.

**Conclusion:** This study found a high burden of hospitalization among chronic disease patients, with drug discontinuation being the leading documented reason. Male sex and regular physical exercise were significantly associated highlighting the need for medication adherence support programs and structured exercise programs integrated into routine chronic disease care, as well as gender-sensitive interventions.

## Introduction

Chronic diseases, also referred to as non-communicable diseases (NCDs), are defined as conditions that persist for one year or more and require ongoing medical attention, limit activities of daily living, or both[1, 2]. These conditions are the leading causes of illness, disability, and death worldwide, accounting for approximately 75% of all non-pandemic-related deaths globally[3]. In 2021 alone, NCDs claimed at least 43 million lives, with 18 million of these deaths occurring before the age of 70 years, of which 82% occurred in low- and middle-income countries[3, 4]. Cardiovascular diseases account for the majority of these deaths (19 million), followed by cancers (10 million), chronic respiratory diseases (4 million), and diabetes (over 2 million). A modifiable risk factors like tobacco use, unhealthy diets, harmful use of alcohol, physical inactivity, and air pollution account for a substantial proportion of this burden [2, 5, 6].

Ethiopia is undergoing a rapid epidemiological transition in which the burden of NCDs is increasing alongside persistent communicable diseases, creating a double burden for an already constrained health system[7]. NCDs have also become an important contributor to hospital-based healthcare utilization. In one Ethiopian study, NCDs accounted for 51.1% of medical admissions, with heart failure reported as the third most common reason for immediate admission[7, 8].

The increasing prevalence of chronic diseases is accompanied by substantial healthcare utilization, especially frequent hospitalization. Patients with chronic diseases may require hospital-based care because of disease complications, treatment failure, acute exacerbations, infections, or medication-related problems [9–11]. In the United States, 20.8% of patients with chronic conditions experienced at least one unplanned hospital admission within one year. Higher Charlson Comorbidity Index scores, as well as conditions such as heart failure, chronic kidney disease, chronic obstructive pulmonary disease, depression, and stroke, were significantly associated with increased risk of hospitalization[10]. In Ethiopia, a study conducted at Hawassa University Comprehensive Specialized Hospital found that 55.2% of outpatients had at least one chronic disease and 17.8% had multimorbidity, highlighting the substantial complexity of chronic disease care beyond individual disease management[12].

Hospitalization among patients with chronic diseases is influenced by a complex interplay of clinical, behavioral, and socioeconomic factors[13]. Multimorbidity is particularly important because the coexistence of several chronic conditions may increase clinical complexity, treatment burden, and the risk of acute deterioration requiring hospital-based care. A study conducted in Bahir Dar, Ethiopia, reported significantly higher odds of hospitalization among patients with three or more chronic NCDs[13]. Behavioral and treatment-related factors may also contribute to hospitalization. Traditional medicine use is common among patients with chronic diseases in Ethiopia, with a pooled prevalence of 56.94%; its use has been associated with factors including female sex, rural residence, longer disease duration, and the presence of complications [14]. Such factors may influence treatment-seeking behavior, continuity of care, and disease management, potentially affecting hospitalization.

Despite the growing burden of chronic diseases and healthcare utilization in Ethiopia, evidence on the burden of hospitalization and its associated factors among patients with multiple chronic diseases remains limited, particularly in the Wolaita Zone. Existing Ethiopian studies have largely focused on individual chronic conditions or specific aspects of healthcare utilization, with limited evidence on hospitalization across patients with multimorbidity. Therefore, this study aimed to determine the burden of hospitalization and associated factors among patients with chronic diseases attending the chronic follow-up clinic at Wolaita Sodo University Comprehensive Specialized Hospital (WSUCSH).

## Methods and participants

### Study setting and Period

The study was conducted at Wolaita Sodo University Comprehensive Specialized Hospital (WSUCSH), located in Wolaita Sodo town, Wolaita Zone, South Ethiopia, located approximately 330 km south of Addis Ababa. WSUCSH serves as a tertiary referral center for chronic disease patients from Wolaita Zone and surrounding areas, providing outpatient chronic follow-up care. Data were collected from November to December 2022 from patients attending the chronic follow-up clinic during their scheduled appointments.

### Study Design

A facility-based cross-sectional study design was employed, using quantitative methods to determine the burden of hospitalization and associated factors among patients with chronic diseases.

### Population

#### Source population

All adult patients with chronic diseases attending the chronic follow-up clinic at WSUCSH during the study period.

#### Study population

All chronic disease patients who came to WSUCSH during the data collection period and fulfilled the inclusion criteria.

### Eligibility Criteria

Patients were eligible for inclusion in the study if they had a confirmed diagnosis of one or more chronic diseases and were on active treatment at the chronic follow-up clinic of WSUCSH. Participants were required to be aged 18 years and above, and to provide written informed consent prior to enrollment. Patients were excluded from the study if they were unwilling to participate, were critically ill at the time of data collection, or had severe psychiatric conditions that would interfere with their ability to provide accurate information. Those with communicable chronic diseases.

### Sample size and Sampling technique

All eligible patients attending the chronic follow-up clinic at WSUCSH, who fulfill inclusion criteria, during the two-month data collection period (November–December 2022) were consecutively recruited. Since chronic disease patients typically visit monthly, each patient’s unique medical record number was recorded at enrollment to prevent double recruitment; only the first encounter per patient was included. A total of 321 patients were enrolled.

### Variables

#### Dependent variable

Hospitalization status (count of any admissions while on treatment).

#### Independent variables

Sociodemographic (age, sex, marital status, religion, ethnicity, education, occupation, income, residence, mode of payment), lifestyle (smoking, alcohol, chat, salt-restricted diet, physical exercise, traditional medicine use), disease-related (type and number of chronic diseases, duration, family history, comorbidity), medication-related (number of medications, polypharmacy, drug allergy, Adverse Drug reaction history), healthcare utilization (missed appointments), and medication adherence.

### Data Collection Procedures

Data were collected using a structured questionnaire administered by trained data collectors and through review of patients’ medical records. Four MSc students in clinical pharmacy on their clinical rotation were used for data collection. The questionnaire was prepared in English, translated to Amharic and Wolayttatto, and back-translated to ensure consistency. Non-compliance to treatment was identified by using the validated Morisky Medication Adherence Scale (MMAS), consisting of 8 items with a dichotomous response (yes/no) with questions asking the patient to respond “yes” or “no” to items 1–7 and a 5-point Likert response for the last item. A positive response indicates a problem with adherence. The total score for each patient is the summation of the scores in each item. Therefore, a score greater than or equal to 3 indicates that the patient’s medication adherence is poor[15].

### Data Quality Assurance

The questionnaire was pre-tested on 5% of the sample at a comparable hospital, and corrections were made before the main study. Data collectors received training on study objectives, procedures, and ethics. The principal investigator supervised data collection daily, and all completed questionnaires were reviewed for completeness.

### Statistical analysis and data management

Data were entered and cleaned in Stata version 17. Descriptive statistics were computed as frequencies and percentages for categorical variables, and as mean ± SD or median (IQR) for continuous variables based on normality (Shapiro-Wilk test). Bivariable analysis used chi-square and independent t-tests. Negative binomial regression was used to identify factors associated with hospital admission rate due to significant overdispersion (LR test of α = 0, p < 0.001). Incidence Rate Ratios (IRRs) with 95% CIs were reported. Variables with p < 0.20 in univariable analysis entered the multivariable model; p <0.05 was considered statistically significant.

### Operational Definitions

*Hospitalization* : any admission while on treatment, including both emergency department and inpatient admissions as documented in the medical record.

*Number of Hospitalizations* ; the total count of hospital admissions (emergency or inpatient) *Polypharmacy* ; concurrent use of five or more medications per day, for a duration of at least one month[16].

*Multimorbidity*; the coexistence of two or more chronic diseases in the same patient, as documented in the medical record[17].

*Traditional Medicine Use*; self-reported use of any traditional or herbal medicine alongside conventional chronic disease treatment

*Missed Appointment*: a scheduled follow-up clinic visits that the patient failed to attend, as reported by the patient or documented in the appointment record

*Body Mass Index (BMI);* weight in kilograms divided by height in meters squared (kg/m²). categorized using CDC and WHO adult cutoffs: underweight (<18.5), normal weight (18.5–24.9), overweight (25.0–29.9), and obese (≥30.0)[18].

### Ethical Considerations

Ethical approval was obtained from the Institutional Review Board of Wolaita Sodo University (CHSM/ERC/02/14). Permission was also obtained from the hospital administration and the head of the diabetes clinic. Written informed consent was obtained from all participants prior to the interview. Patient confidentiality was maintained throughout the study, and all data were anonymized prior to analysis. Participants were informed of their right to withdraw from the study at any time without affecting their medical care.

## Results

### Sociodemographic characteristics of study participants

The majority of the 321 study participants were female (54%), urban residents (67%), and married/living together (77%). The mean age was 46.46 ± 13.62 years, with the largest age group being 46–64 years (45%), followed by 25–45 years (38%). Regarding education, more than one-third (37%) had completed 9–12 grade, and 19.9% had a degree or above. The most common occupations were merchant (30%) and employee (25%), while farmers accounted for 18%. Monthly income was distributed almost equally across low, medium, and high tertiles. Nearly half (49.2%) had health insurance coverage, and 35.5% paid out-of-pocket (Table 1).

**Table 1.** Sociodemographic characteristics of adult patients with chronic diseases attending the chronic follow-up clinic at WSUCSH (n = 321)

| Variable | Category | Frequency (n) | Percentage (%) |
| --- | --- | --- | --- |
| Sex | Female | 174 | 54.2 |
|  | Male | 147 | 45.8 |
| Age (years) | Mean ± SD | 46.46 ± 13.62 | — |
|  | Range | 15 – 80 | — |
| Age group (years) | <25 | 19 | 5.9 |
|  | 25–45 | 123 | 38.3 |
|  | 46–64 | 145 | 45.2 |
| | ≥65 | 34 | 10.6 |
| Marital status | Married, living together | 246 | 76.6 |
|  | Single | 38 | 11.8 |
|  | Married, not living together | 18 | 5.6 |
|  | Divorced | 12 | 3.7 |
|  | Widowed | 7 | 2.2 |
| Educational status | 9–12 grade | 118 | 36.8 |
|  | Degree and above | 64 | 19.9 |
|  | 1–8 grade | 44 | 13.7 |
|  | Diploma | 44 | 13.7 |
|  | Read and write only | 39 | 12.1 |
|  | Unable to read & write | 12 | 3.7 |
| Occupation | Merchant | 96 | 29.9 |
|  | Employee | 79 | 24.6 |
|  | Farmer | 59 | 18.4 |
|  | Other | 58 | 18.1 |
|  | Driver | 29 | 9.0 |
| Monthly income (ETB) | Low (0–1,500) | 112 | 34.9 |
|  | Medium (1,600–4,000) | 102 | 31.8 |
|  | High (4,500–18,000) | 107 | 33.3 |
| Residence | Urban | 216 | 67.3 |
|  | Rural | 105 | 32.7 |
| Mode of payment | Insurance | 158 | 49.2 |
|  | Pocket/Self-pay | 114 | 35.5 |
|  | NGO | 49 | 15.3 |

➤ Age is reported as mean ± SD because it was approximately normally distributed (Shapiro-Wilk $p = 0.079$ ).

### Lifestyle characteristics of study participants

Smoking was rare (0.9% current; 4.0% former), and only 12.8% reported current alcohol use. Chat use was reported by 5.3%. Adherence to salt-restricted diet was suboptimal 43.3% reported "rarely" and only 31.2% exercised regularly, while 37.7% did not exercise at all. The median BMI was 23 (IQR: 20–25), and the range was 11–34. Table 2

**Table 2.** Lifestyle characteristics of adult patients with chronic diseases attending the chronic follow-up clinic at WSUCSH (n = 321)

| *Variable* | *Category* | *Frequency (n)* | *Percentage (%)* |
| --- | --- | --- | --- |
| Smoking habit | No | 305 | 95.0 |
|  | Used to | 13 | 4.0 |
|  | Yes | 3 | 0.9 |
| Alcohol use | No | 224 | 69.8 |
|  | Used to | 56 | 17.4 |
|  | Yes | 41 | 12.8 |
| Chat use | No | 297 | 92.5 |
|  | Used to | 7 | 2.2 |
|  | Yes | 17 | 5.3 |
| Salt-restricted diet | Always | 64 | 19.9 |
|  | Usually | 90 | 28.0 |
|  | Rarely | 139 | 43.3 |
|  | Never | 28 | 8.7 |
| **regular physical exercise** | No | 119 | 37.1 |
|  | Regularly | 92 | 28.7 |
|  | Sometimes | 110 | 34.3 |
| **BMI (kg/m<sup>2</sup>)\*** | Median (IQR) | 23 (20–25) | — |
|  | Range | 11 – 34 | — |

\*-BMI is reported as median (IQR) because of significant right skewness (Shapiro-Wilk $p < 0.001$ ).

Regarding to MBI of study participants, based on WHO classification, about half of them (50.5%) had normal weight, and 3.1% were obese (Fig 1).

**Fig 1.**
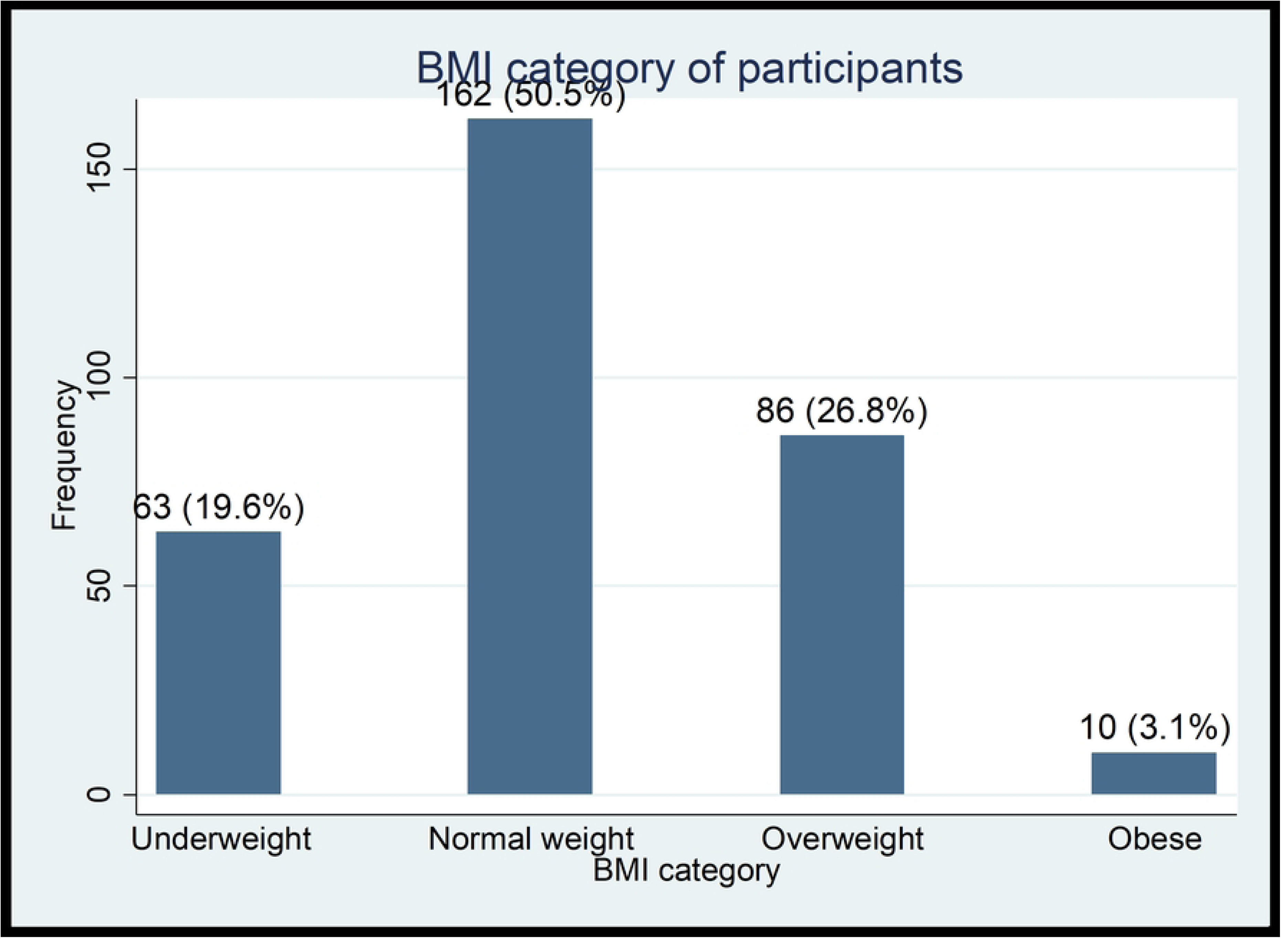
BMI distribution among adult patients with chronic diseases attending the chronic follow-up clinic at WSUCSH (n = 321)

### Disease and Medication related characteristics

Among the 321 patients studied, the mean number of chronic diseases was 2.40 ± 1.02 per patient. Multimorbidity is highly prevalent, with the majority (59.8%) had 2 to 3 chronic conditions (Fig 2).

**Fig 2.**
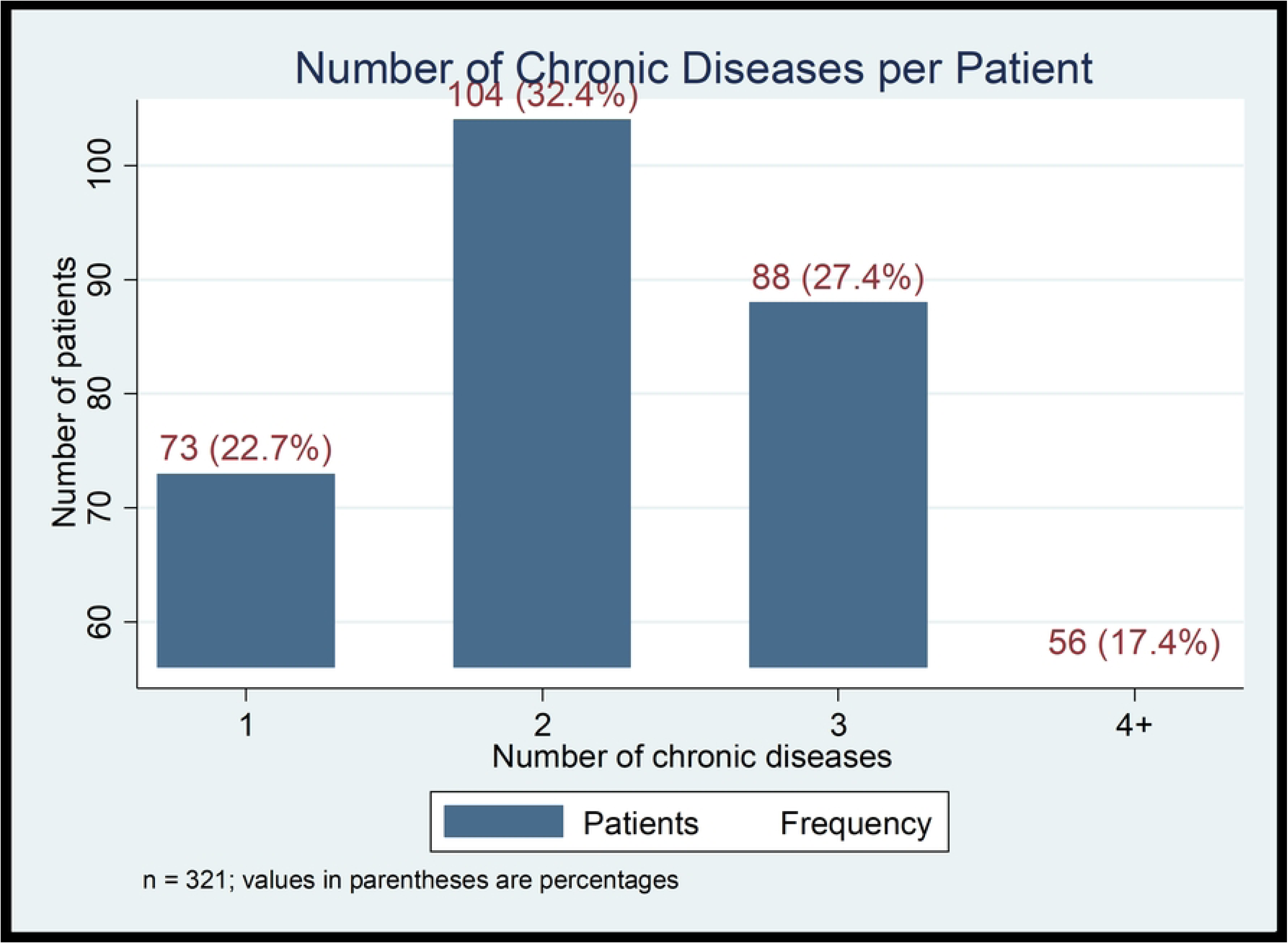
Distribution of chronic diseases among adult patients with chronic diseases attending the chronic follow-up clinic at WSUCSH (n = 321)

Out of 321 patients with chronic diseases, half of them,159 (49.5%) had at least one hospitalization while on treatment. Among the 159 hospitalized patients, the most common reason was drug discontinuation (56, 35.2%), followed by disease complication (41, 25.8%) and infection (31, 19.5%). The mean number of hospitalizations was 0.97 ± 1.20, with a range of 0–6 Table 3.

**Table 3.**
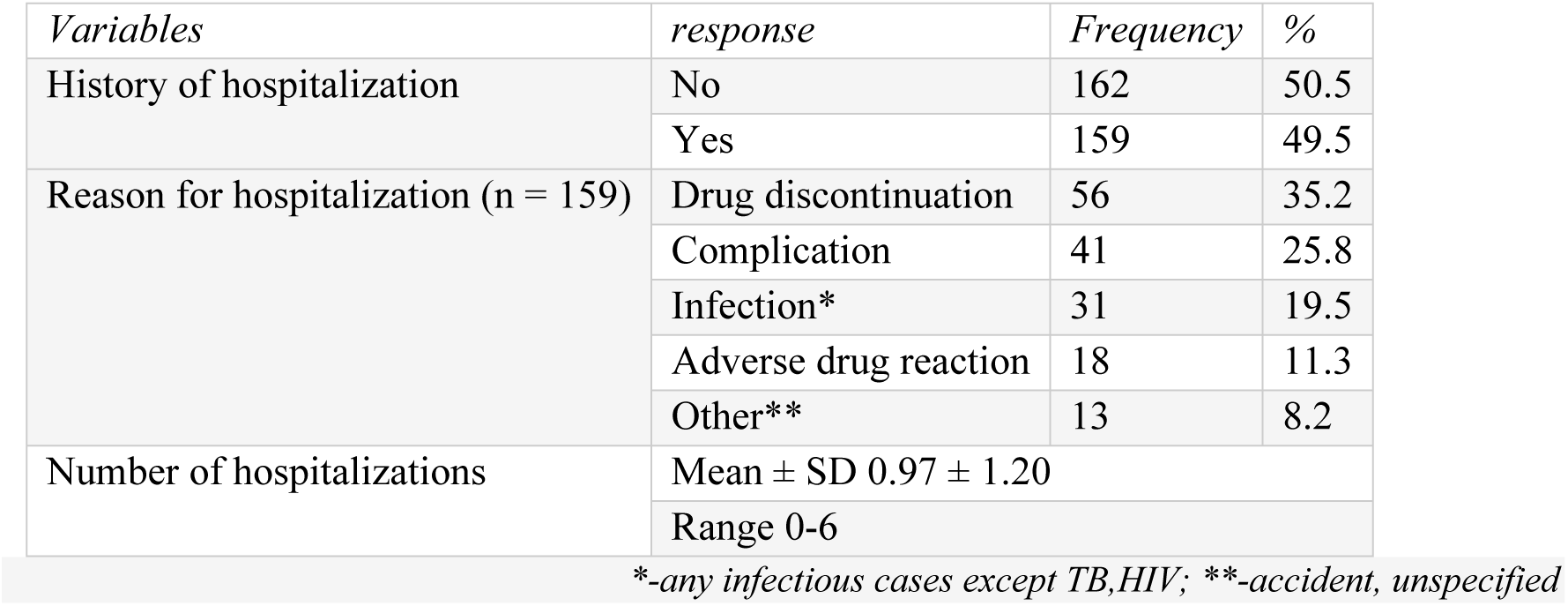
hospitalization status of adult patients with chronic diseases attending the chronic follow-up clinic at WSUCSH (n = 321)

**Table 4.**
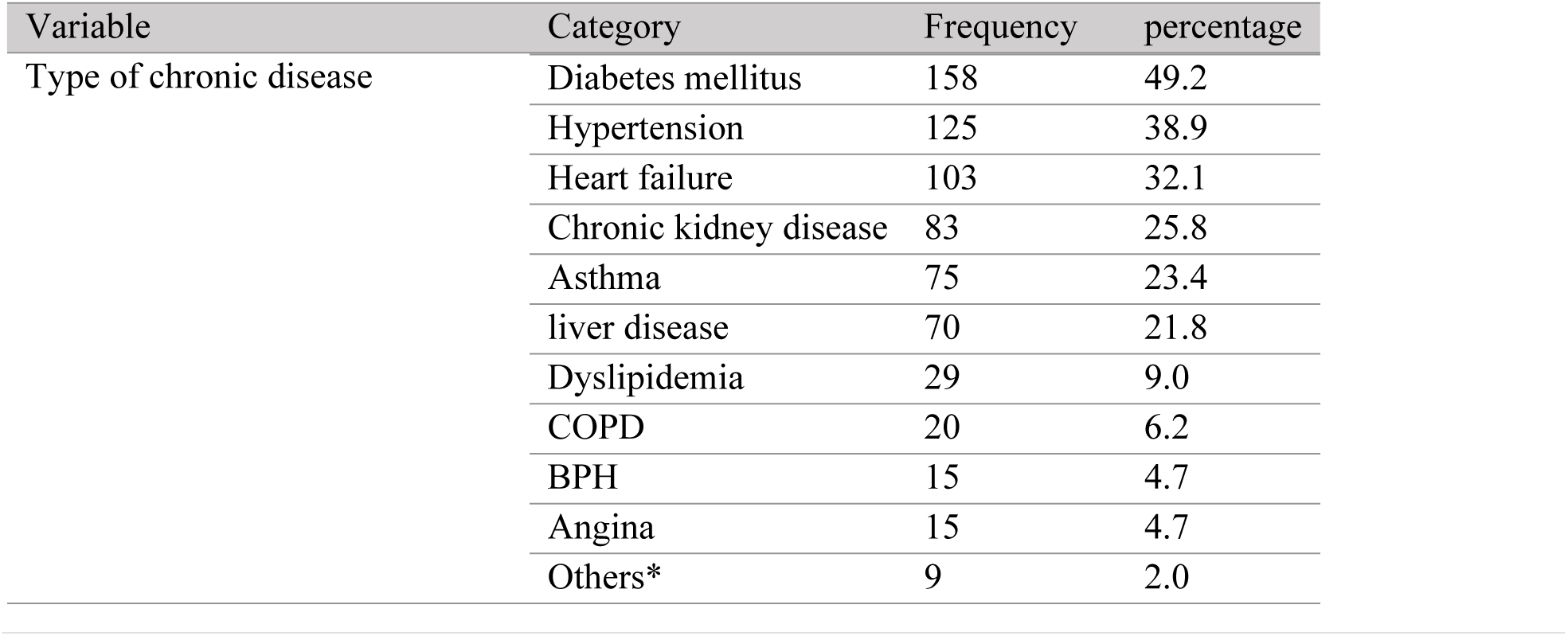
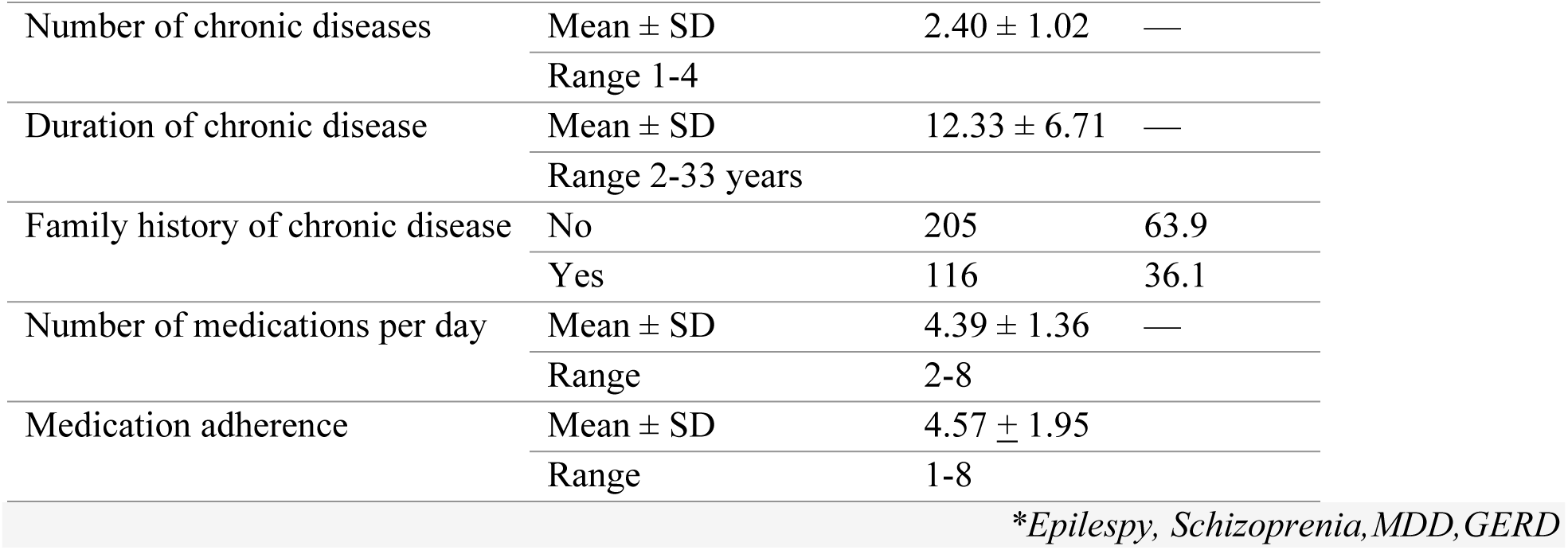
Disease and medication related characteristics of adult patients with chronic diseases attending the chronic follow-up clinic at WSUCSH (n = 321)

**Table 5.**
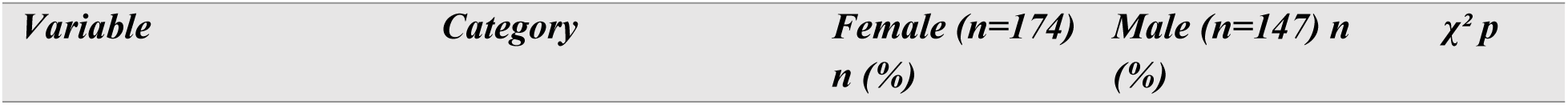
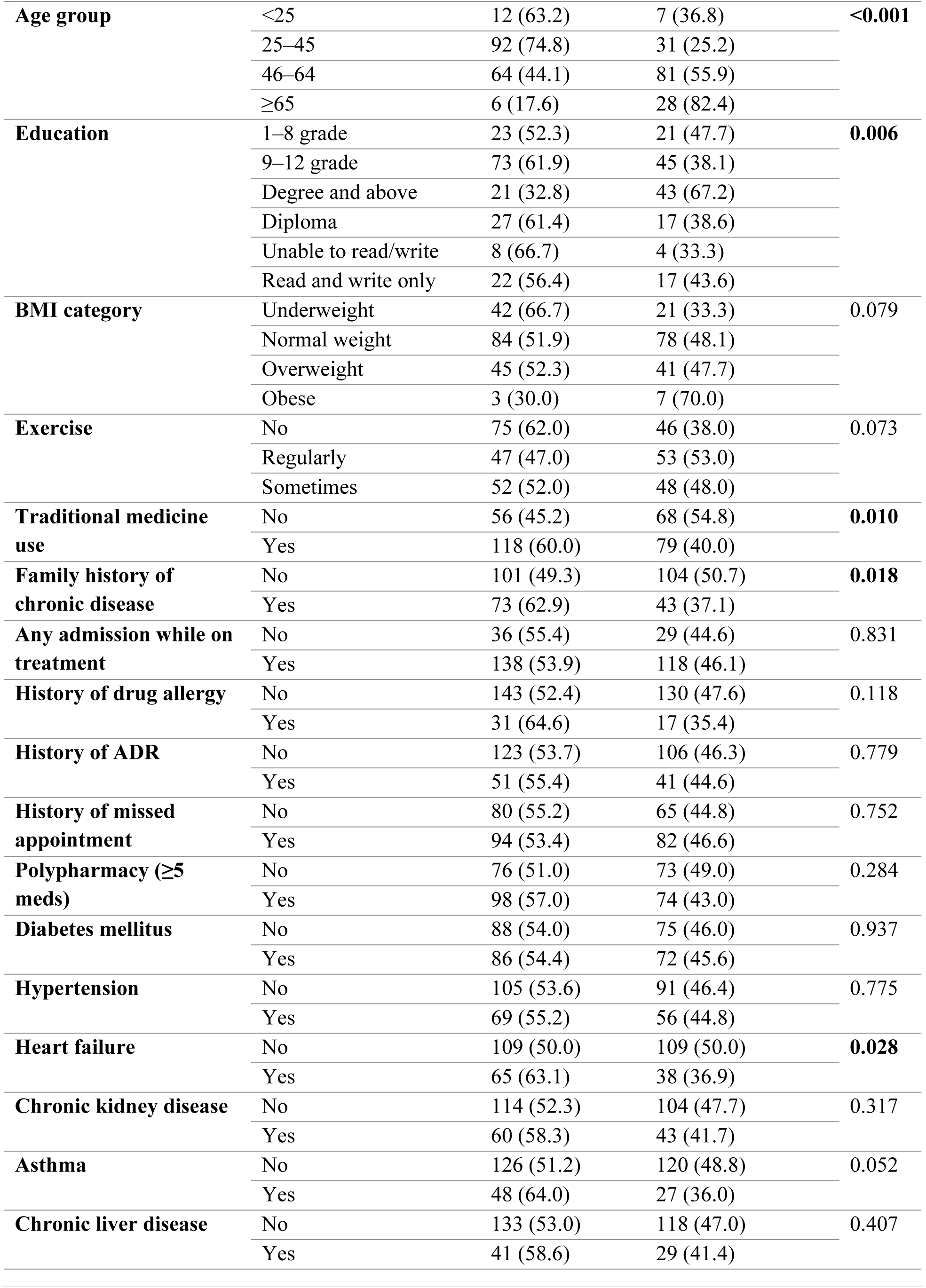
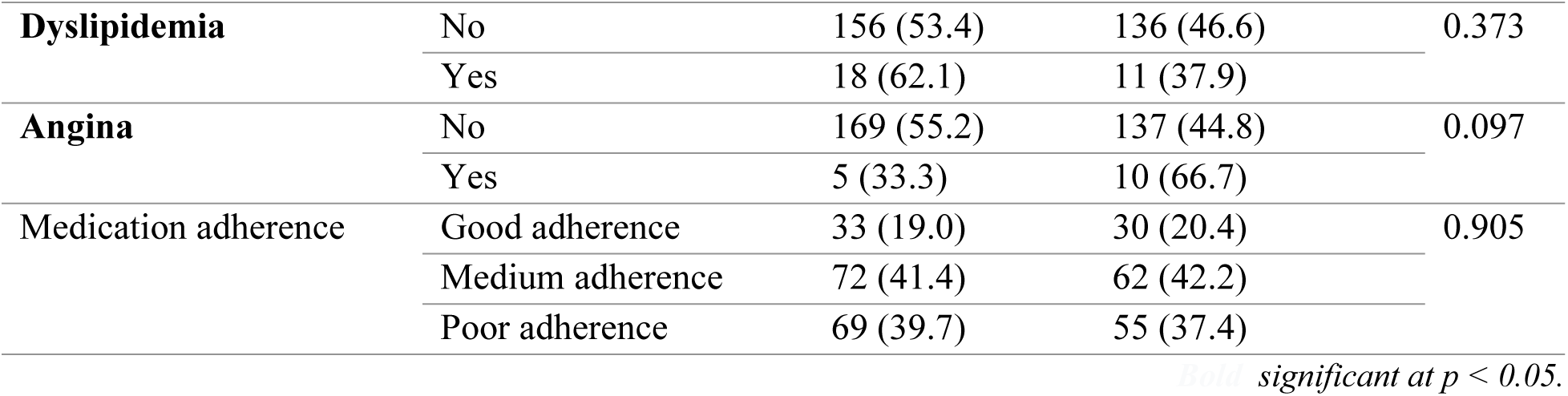
Sex differences of categorical variables of adult patients with chronic diseases attending the chronic follow-up clinic at WSUCSH (n = 321)

**Table 6.** Sex differences of continuous variables of adult patients with chronic diseases attending the chronic follow-up clinic at WSUCSH (n = 321)

| Variable | Female Mean ± SD | Male Mean ± SD | Mean difference (Female – Male) | 95% CI | p-value |
| --- | --- | --- | --- | --- | --- |
| Age (years) | $42.16 \pm 11.87$ | $51.54 \pm 13.85$ | -9.38 | -12.21 to -6.56 | **<0.001** |
| BMI (kg/m<sup>2</sup>) | $21.80 \pm 4.40$ | $22.91 \pm 3.92$ | -1.11 | -2.03 to -0.18 | **0.019** |
| Number of hospitalizations | $1.48 \pm 1.65$ | $1.97 \pm 1.96$ | -0.48 | -0.88 to -0.09 | **0.017** |
| Monthly income (ETB) | $2,599 \pm 2,931$ | $5,482 \pm 4,325$ | -2,882 | -3,684 to -2,081 | **<0.001** |
| Number of medications/days | $4.49 \pm 1.38$ | $4.27 \pm 1.32$ | 0.22 | -0.08 to 0.52 | 0.142 |
| Number of chronic diseases | $2.43 \pm 0.96$ | $2.35 \pm 1.09$ | 0.08 | -0.15 to 0.30 | 0.501 |
| Duration of disease (years) | $11.92 \pm 6.43$ | $12.82 \pm 7.03$ | -0.90 | -2.38 to 0.57 | 0.230 |

**Table 7.** Factors Associated with Hospitalization of adult patients with chronic diseases attending the chronic follow-up clinic at WSUCSH.

| *Variable* | *IRR* | *Std. Err.* | *z* | *p-value* | *95% CI* |
| --- | --- | --- | --- | --- | --- |
| **Gender** |  |  |  |  |  |
| Male (vs Female) | 1.4355 | 0.1752 | 2.96 | **0.003** | **1.1302 – 1.8233** |
| **Exercise** |  |  |  |  |  |
| Regularly (vs No) | 0.4965 | 0.0782 | −4.44 | **0.000** | **0.3645 – 0.6762** |
| Sometimes (vs No) | 1.0799 | 0.1495 | 0.56 | 0.579 | 0.8233 – 1.4164 |
| Constant | 1.6484 | 0.1763 | 4.67 | 0.000 | 1.3367 – 2.0330 |
| /lnalpha | −0.6114 | 0.1957 |  |  | −0.9949 – −0.2279 |
| alpha | 0.5426 | 0.1062 |  |  | 0.3697 – 0.7962 |
| *Model fit: Observations 321; LR $\chi^2(3)$ 31.46; Prob > $\chi^2$ 0.0000; Pseudo R<sup>2</sup> 0.0276 ; LR test of alpha = 0; $\chi^2(1)$ = 64.68; $p < 0.001$* |  |  |  |  |  |

*Note: IRR = incidence rate ratio; CI = confidence interval. Reference categories: gender = Female; exercise = No regular exercise.*

Diabetes mellitus (49.2%) and hypertension (38.9%) were the most prevalent conditions. The mean number of chronic diseases was 2.40 ± 1.02. Polypharmacy was present in 53.6%. Over half (54.8%) had missed appointments, and 61.4% reported using traditional medicine.

Regarding medication adherence, the mean MMAS-8 score was 4.57 (SD = 1.95; range 1–8), 124 (38.6%) had poor adherence and 63 (19.6%) had good adherence (score 8). Overall, 80.4% of participants had less than good adherence, indicating that medication adherence was suboptimal (Fig 3).

**Fig 3:**
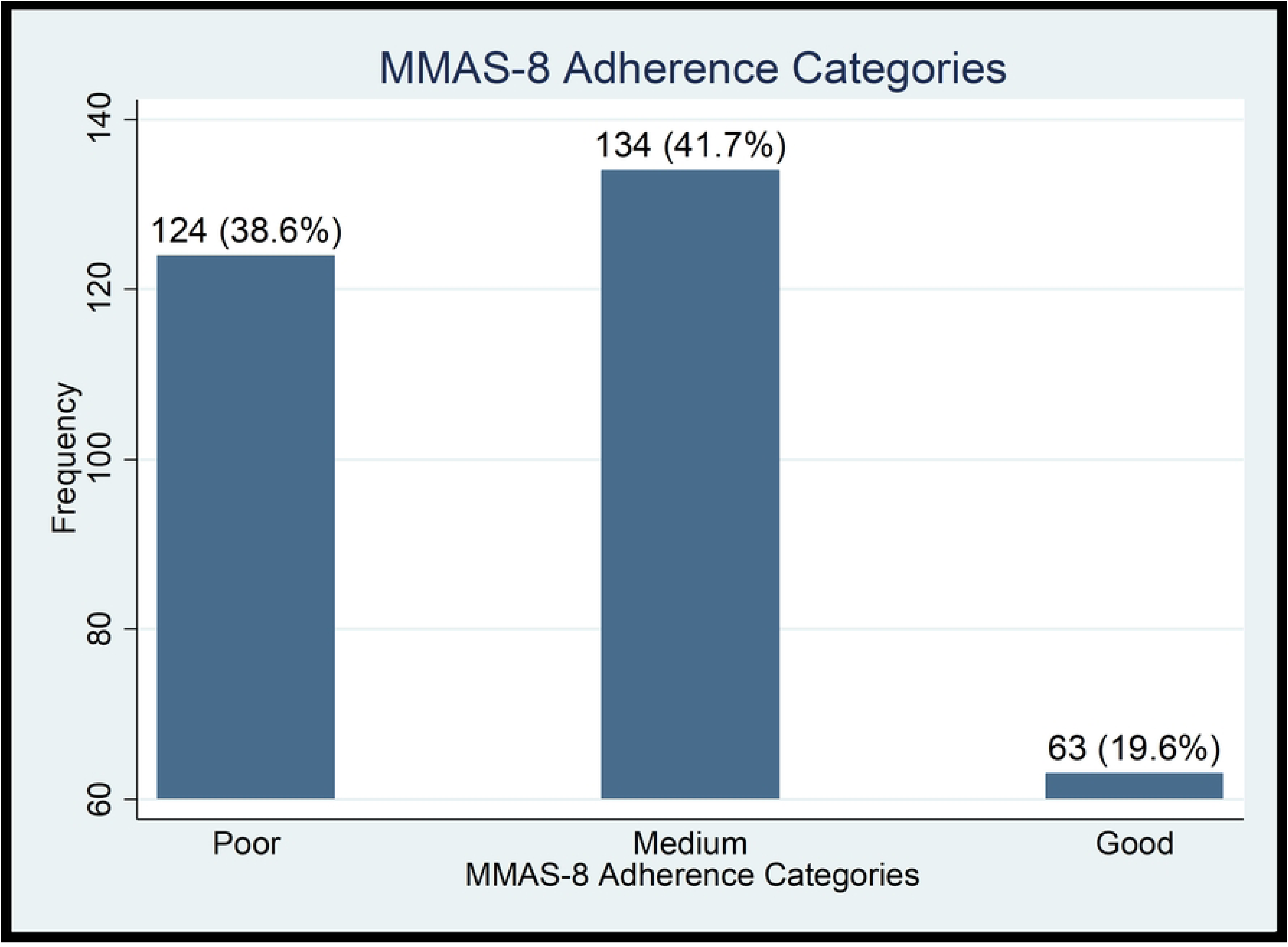
Medication adherence among adult patients with chronic diseases attending the chronic follow-up clinic at WSUCSH (n = 321)

### Sex Differences among study variables

Significant sex differences were found for age group, education, traditional medicine use, family history of chronic disease and heart failure(all p < 0.05). Females were more likely to use traditional medicine (60% vs 40%), report family history (63% vs 37%), and have heart failure (63% vs 37%). Males had significantly higher mean age, BMI, income, and number of admissions(table-6). No significant sex differences were observed for diabetes, hypertension, CKD, CLD, dyslipidemia, COPD, polypharmacy, medication adherence, history of ADR, missed appointments, or overall admission history. Table-5

Continuous variables were compared between sexes using independent-sample t-tests. Male patients were significantly older (51.54 ± 13.85 vs 42.16 ± 11.87 years; p < 0.001), had higher BMI (22.91 ± 3.92 vs 21.80 ± 4.40 kg/m²; p = 0.019), more hospital admissions (1.97 ± 1.96 vs 1.48 ± 1.65; p = 0.017), and higher monthly income (5,482 ± 4,325 vs 2,599 ± 2,931 ETB; p < 0.001) compared with female patients. Table-6

### Factors Associated with Hospitalization

In the final multivariable negative binomial regression model, two variables were significantly associated with hospitalization of patients with chronic diseases at WSUCSH. Male patients had 44% more hospital admissions than female patients (adjusted IRR = 1.44; 95% CI: 1.13–1.82; p = 0.003). Regular physical exercise was significantly associated with fewer hospitalizations: patients who exercised regularly had 50% fewer hospitalization than those who did not exercise (adjusted IRR = 0.50; 95% CI: 0.36–0.68; p < 0.001). Table-7

## Discussion

This study assessed the burden and factors associated with hospitalization among 321 patients with chronic diseases attending the chronic follow-up clinic at WSUCSH, South Ethiopia. The study found that 66% of patients reported at least one hospital admission while on treatment, with a mean of 1.70 ± 1.81 admissions per patient. The high burden of hospitalization observed in this study, is considerably higher than the 9.4% reported in a longitudinal study in Bahir Dar, Northwest Ethiopia[13], and in Eastern Ethiopia, 9.85 per 1000 person-years[19]. These discrepancies are can be explained by differences in the definition of hospitalization and the observation period. Our study employed a broader definition of hospitalization including both emergency department visits and inpatient admissions over the entire treatment period whereas previous studies restricted their outcome to inpatient admissions within a fixed follow-up window. However, the current findings are consistent with the study by Noor et al. in Sudan[20], which showed that 71.8% of medical admissions in a Sudanese tertiary hospital were attributable to NCDs.

The finding that drug discontinuation was the leading documented reason for hospitalization is consistent with study at Felege Hiwot Comprehensive Specialized Hospital in Northwest Ethiopia [21] and Tikur Anbessa Specialized Hospital in Addis Ababa [8]. The Longman et al. study in rural Australia similarly identified that patients who reported forgetting to take their medications were more likely to be admitted frequently, reinforcing the link between medication management and hospitalization risk[22].

The significant association between male sex and higher hospitalization rate in our study is corroborated by previous studies like, a study at Saint Paul’s Hospital in Addis Ababa [23], and at Hawassa University Comprehensive Specialized Hospital[12]. A large-scale analysis of 105,025 medical records in rural Ethiopia also reported that men had more admissions for leading causes[24]. The Longman et al. study in rural Australia[22] also found that being male was associated with frequent admission among older patients with chronic disease, consistent with our findings. A retrospective multicenter study of 18,448 patients with multiple chronic conditions in the United States found that male sex was not significantly associated with unplanned admissions after adjustment, suggesting that the effect of sex may be context-specific and influenced by differences in healthcare access and disease patterns[10]. The higher hospitalization rate among males in our study may reflect later health-seeking behavior, greater disease severity at presentation, and lower adherence to follow-up care among men, as suggested by qualitative research from Kenya which found that men often delay seeking care due to traditional masculine norms and perceive hospital visits as a last resort[25].

The finding that regular physical exercise was associated with a lower number of hospitalizations is consistent with a large body of international evidence from longitudinal and interventional studies, though the cross-sectional design of our study precludes causal inference. A Cochrane review of 107 trials involving 26,886 people with coronary heart disease demonstrated that exercise-based cardiac rehabilitation reduces all-cause hospital admissions by 35% and the risk of myocardial infarction by 28%[26]. A nationwide population-based study in Korea found that a 500 MET-min/week increase in physical activity was associated with an 8% reduction in all-cause hospitalization and a 13% reduction in respiratory hospitalization among patients with COPD[25] [27]. A UK Biobank study of accelerometer-measured physical activity and hospitalization risk across 25 common health conditions found that increasing moderate-to-vigorous physical activity by 20 minutes per day was associated with reductions in hospitalization ranging from 3.8% across conditions[28]. The Longman et al. study in rural Australia found that health behaviour, including physical activity, were among the potentially modifiable factors that could reduce hospital admissions[22]. The Previous study in Hawassa also found that patients who exercised were significantly less likely to have multimorbidity[12]. Our finding that occasional exercise showed no significant association with hospitalization rate further may suggest that consistency of physical activity may be required to observe any association with clinical outcomes. This has important implications for chronic disease management programs in Ethiopia, where structured and regular exercise programs remain rare and physical inactivity is highly prevalent, with studies showing that nearly one-third of patients are unable to perform physical activity due to chronic medical problems and aging[27]. However, because our study was cross-sectional and exercise was assessed by self-report, reverse causation cannot be excluded, patients with fewer hospitalizations and better-controlled disease may be more capable of engaging in regular exercise, and longitudinal studies are needed to clarify the direction of this association.

The findings for polypharmacy, multimorbidity, and medication adherence in our study contrast with some previous research like, a study in Bahir Dar[13]. The lack of significant associations in our study may reflect differences in study design, the broad definition of admission used, or the high baseline burden of disease in our cohort (mean 2.40 chronic diseases) reducing the contrast needed to detect differences.

The high prevalence of traditional medicine use in our cohort is consistent with the pooled estimate reported in a systematic review and meta-analysis of herbal medicine use among patients with chronic disease in Ethiopia[14]. Our finding that females were significantly more likely to use traditional medicine aligns with these findings. Similarly, other studies on herbal medicine use among diabetes patients in Ethiopia reported that nearly half of participants used herbal medicine[14, 29].

## Limitation of the study

This study provides a comprehensive assessment of hospitalization burden across multiple chronic diseases in a tertiary hospital in southern Ethiopia, addressing a gap in the literature. However, the following limitations should be acknowledged. The cross-sectional design precludes causal inference, and the single tertiary referral setting may limit generalizability to primary care or other regions. Hospitalization history and reasons relied on self-report and medical records, which are susceptible to recall and documentation bias. The sample size was determined by consecutive recruitment over two months rather than an a priori calculation, and although post-hoc power was adequate, some associations may have been underpowered. Physical exercise was self-reported without objective measurement, and reverse causation cannot be excluded. Reasons for hospitalization were categorized using a priority system that may not capture multi-causal admissions, potentially underestimating the full burden of acute care utilization.

## Conclusion

This study found multimorbidity was highly prevalent, with the majority of patients having 2 to 3 chronic conditions. The study also revealed a high burden of hospitalization, with approximately two-thirds of patients reporting at least one hospitalization while on treatment. Drug discontinuation was the leading documented reason for hospitalization, followed by disease complications and infections. Male sex was independently associated with a higher hospitalization rate, whereas regular physical exercise was associated with fewer hospitalizations. These findings suggest that hospitalization among chronic disease patients in this setting is primarily associated with modifiable behavioral factors, specifically medication discontinuation and physical inactivity as well as male sex, which may potentially reflect delayed health-seeking behavior.

## Recommendations

Healthcare providers should counsel on medication adherence, explore reasons for discontinuation, prescribe regular exercise, and actively engage male patients in timely follow-up. Policymakers should support structured exercise programs and develop gender-sensitive strategies that encourage earlier healthcare engagement among men; and finally, researchers should conduct longitudinal studies on exercise and male sex as predictors of hospitalization, qualitative research on drug discontinuation and male health-seeking behavior, and intervention trials testing adherence and exercise programs.

## Data Availability

The datasets used and/or analyzed during the current study are available from the corresponding author on reasonable request

## Acknowledgments

We are indebted to Wolaita Sodo University Hospital, data collectors and to the participants of this study for providing us with their valuable time, information, help and cooperation.

## Declarations

### Author Contributions

*Conceptualization*: Hailu Chare, Yalemzerf Worku, Zewde Zema, Zinabu Desalegn

*Data curation*: Hailu Chare, Mengistu Meskele, Yalemzerf Worku

*Formal analysis*: Hailu Chare, Mengistu Meskele

*Investigation*: Hailu Chare, Yalemzerf Worku, Zinabu Desalegn

*Methodology*: Hailu Chare, Mengistu Meskele,Yalemzerf Worku, Zewde Zema, Zinabu Desalegn

*Supervision*: Zewde Zema, Zinabu Desalegn, Mengistu Meskele

*Writing – original draft*: Hailu Chare

*Writing – review & editing*: Hailu Chare, Mengistu Meskele,Yalemzerf Worku, Zewde Zema, Zinabu Desalegn

### Generative artificial intelligence

None used

## Acronyms

[ADR]: Adverse Drug reaction,
[BMI]: Body Mass Index;
[CKD]: Chronic Kidney Disease,
[COPD]: Chronic Obstructive Pulmonary Disease,
[CDC]: Center for Disease Control,
[CLD]: Chronic Liver disease,
[DM]: Diabetes Mellitus,
[HF]: Heart Failure,
[NCDs]: Non-communicable Diseases,
[T2DM]: Type-2 Diabetes Mellitus,
WHO: World Health Organization,
[WSUCSH]: Wolaita Sodo University Comprehensive Specialized Hospital

